# The association of ABO blood group with the asymptomatic COVID-19 cases in India

**DOI:** 10.1101/2021.04.01.21254681

**Authors:** Prajjval Pratap Singh, Abhishek K Srivastava, Sudhir K Upadhyay, Ashish Singh, Pranav Gupta, Sanjeev Maurya, Shashank Upadhyay, Rudra Kumar Pandey, Anshika Srivastava, Priya Dev, Vanya Singh, Rahul Mishra, Manoj Kumar Shukla, Govind Chaubey, Pradeep Kumar, Vandana Rai, Yamini B Tripathi, Abhishek Pathak, Vijay Nath Mishra, Chandana Basu Mallick, Pankaj Shrivastava, Gyaneshwer Chaubey

## Abstract

The COVID-19 pandemic has resulted several waves of infection in many countries worldwide. The large variations in case fatality ratio among different geographical regions suggests that the human susceptibility against this virus varies substantially. Several studies from different parts of the world showed a significant association of ABO blood group and COVID-19 susceptibility. It was shown that individuals with blood group O are at the lower risk of coronavirus infection. To establish the association of ABO blood group in SARS-CoV-2 susceptibility, we for the first time analysed SARS-CoV-2 neutralising antibodies as well as blood groups among 509 random individuals from three major districts of Eastern Uttar Pradesh region of India.. Interestingly, we found neutralising antibodies in significantly higher percentage of people with blood group AB (0.36) followed by B (0.31), A (0.22) and lowest in people with blood group O (0.11). This indicates that people with blood group AB are at comparatively higher risk of infection than other blood groups. Further, in line to previous reports we too observed that people with blood group O have significantly decreased risk of SARS-CoV-2 infection. Thus, among the asymptomatic SARS-CoV-2 infected individuals with blood group AB has highest, whilst blood group O has lowest risk of infection.

## Introduction

COVID-19 has impacted life of billions because of its detrimental nature. With extensive and ongoing research, we are slowly understating the nature of this novel SARS-CoV-2 virus first time transmitted to the humans ^1–6^. With the growing knowledge about this disease, it is clear that there are certain risk factors associated with morbidity and mortality ^7–9^. More importantly, several of the studies have found strong association of the ABO blood group and COVID-19 with morbidity and mortality ^6,10–14^. In the past, there have been several studies on the association of ABO blood group with the diseases. For example, individual with the blood group O were reported to be more susceptible to the Cholera in Gangetic plain populations ^15^ and *Helicobacter pylori* infection^16^. However, blood group O was found to be less susceptible for Dengue ^17,18^ and SARS (Severe Acute Respiratory Syndrome) viruses ^14,19^.

The ABO blood type is administered by the gene *ABO*, present at chromosome 9 ^20^. Studies have found that the this gene modulates the COVID-19 susceptibility directly or indirectly ^21–23^. Several variants of this gene affect morbidity and mortality in COVID-19 and many other diseases. For example, it affects red blood related physiology ^24,25^, venous thromboembolism ^26^, type 2 diabetes ^27^, ischemic stroke ^28^, heart related functions ^29^ and coronary artery disease ^29–31^. Thus, investigating the association of ABO blood type with SARS-CoV-2 infection, it is feasible to ascertain the factors affecting the susceptibility against SARS-CoV-2 in humans.

In the present study, we sought to investigate the association between asymptomatic COVID-19 positive people with their blood types using random serosurvillance and blood group testing of street vendors in northern India.

## Materials and Method

In our survey, we used two commercially available kits Coviscreen™ and ERYCARD™ 2.0 to determine the seropositivity and the blood groups respectively. Theses kits were kind gift by Biosense Technologies, India. Similar to our previous work ^32^, we focussed on the urban populations in our survey. A total of 509 healthy individuals were screened for both the tests. The participants were between the age of 18-65 years from the three districts of the Eastern Uttar Pradesh state (Supplementary Fig. 1). We focussed on the urban healthy vendors who have neither been diagnosed with COVID-19, nor had been sick with any associated symptoms in the recent past. We excluded those individuals whose family members had ever been diagnosed with COVID-19. This study was conducted between months of September 2020 to October 2020. The aim of the study was explained to the people and informed consent were obtained. This study was approved by the ethical committee of Banaras Hindu University, Varanasi, India.

Both COVID-19 and blood group testing were done using manufacturer’s instructions provided in the kit. A sample test of Coviscreen™ and ERYCARD™ 2.0 has been shown in Supplementary Fig. 2. The frequency of seropositive and blood groups were calculated (Supplementary Table 1). The frequency bar plot of each blood group was drawn with 95% CI (Figure 1). We have also collected blood group data of the same region for comparison from published sources ^33–40^.

**Figure 1.**
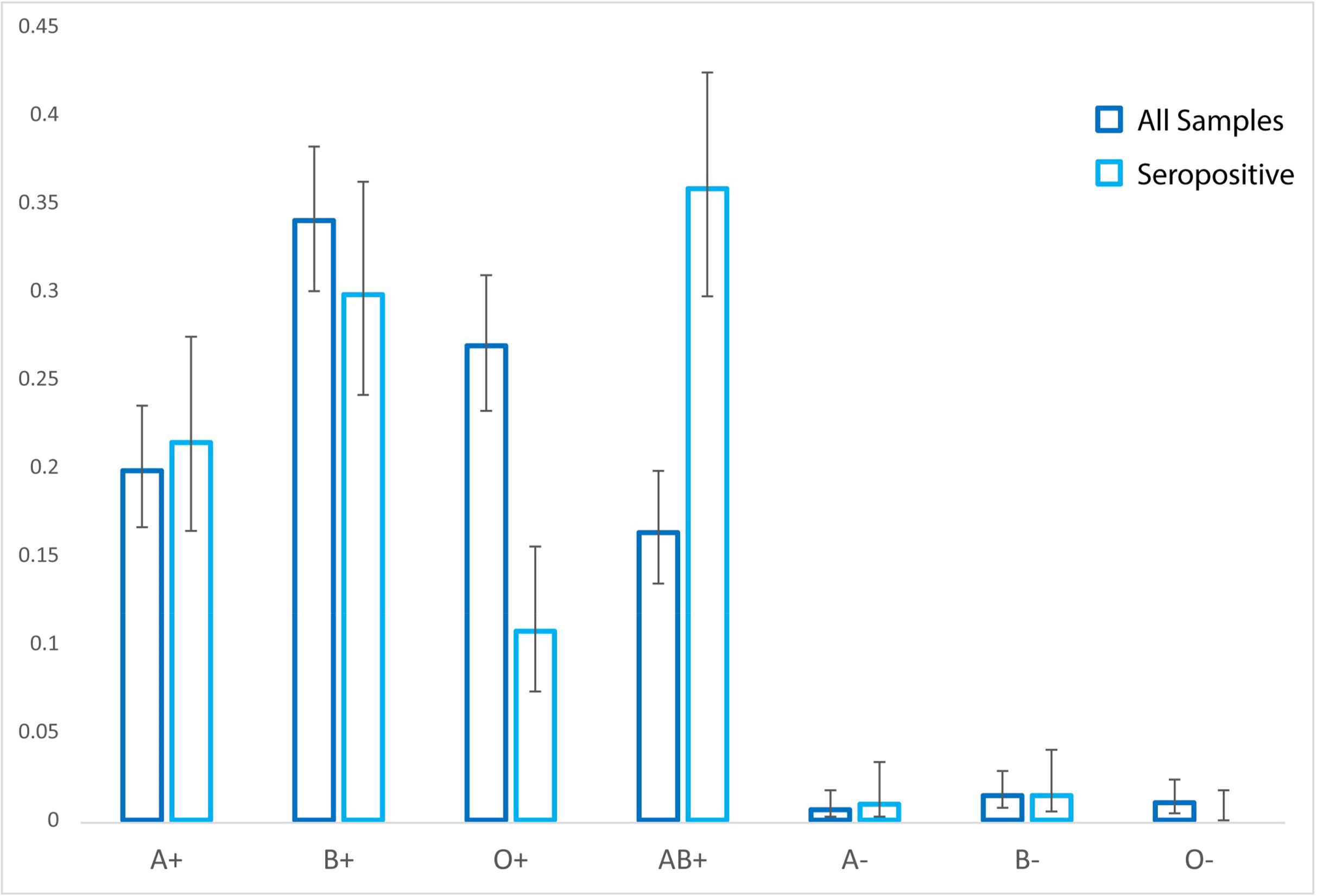
The comparative bar-plot of various blood groups among studied groups. The error bars have been calculated at 95% confidence interval.

The data was analysed using two tailed Chi square test. Odds ratio (OR) and 95% Confidence interval (CI) were calculated (Table 1 and Supplementary Table 2). Statistical computations were performed using SPSS (ver. 25).

**Table 1.** Various statistical tests on our data to infer the association and risk of ABO blood group for COVID-19. OR= Odds Ratio, CI= Confidence interval

|  | A | B | AB | O |
| --- | --- | --- | --- | --- |
| Chi square | 0.632 | 2.564 | 101.331 | 53.276 |
| p value | 0.427 | 0.109 | <0.001 | <0.001 |
| OR | 1.222 | 0.725 | 26.783 | 0.176 |
| 95% CI | 0.792 - 1.884 | 0.500 - 1.052 | 11.3888 - 62.986 | 10.108 - 0.288 |

## Results and discussion

We collected data of seroprevalence as well as blood group affiliation among 509 individuals from three districts of Eastern Uttar Pradesh region (Supplementary Table 1). The seroprevalence among all the studied districts was >0.4. The high level of seroprevalence among these districts suggests a hidden undercurrent wave of infection mainly driven by asymptomatic individuals. It is imperative to stress that, this is not the story of a particular region of India. Remarkably, other independent regions in the country have also demonstrated a high level of seroprevalence ^41,42^. Nevertheless, it is not uniform, rather sporadic.

In the collection of random 509 samples, we found a frequency distribution of 0.204, 0.354, 0.279 and 0.163 for blood groups A, B, O and AB respectively (Supplementary Table 1). Blood group B is most common blood group followed by blood group O, whereas blood group AB is least common among the populations of Eastern Uttar Pradesh region. To test if our sampling covered a true blood group distribution of this region, we have collected the published data of the same region ^33–40^ and performed a regression analysis. The adjusted R square value (96.7% ±2.7%) showed a significantly high level of correlation between our data as well as published data, suggesting that our sampling indeed captured the regional distribution of the blood groups.

We further grouped COVID-19 positive samples and estimated their ABO blood group division. The 215 seropositive individuals showed their ABO blood group distribution of 0.223, 0.312, 0.107 and 0.358 for A, B, O and AB blood groups respectively (Supplementary Table 1). The relative comparison of blood group distribution have shown large discrepancy for the blood groups O and AB (Fig. 1). Blood group AB was significantly higher, whereas blood group O was significantly lower among seropositive group (p<0.001) (Table 1). This result was also consistent with the comparison of seropositive individuals with the published data (Supplementary Fig. 3 and Supplementary Table 1). This suggested strong association of blood groups O and AB with COVID-19 susceptibility. The risk estimation revealed several fold higher risk of infection to blood group AB and lower risk for blood group O type (Table 1 and Supplementary Table 2). On the risk scale, our investigation suggested blood group AB at the maximum, followed by blood group A and B, whereas blood group O had lowest.

We would like to stress that our data does not include COVID-19 severe patients, due to our sampling methodology. Thus, it limits us to understand the association of blood group with the COVID-19 severity. On the other hand, it is highly enriched for the asymptomatic patients. Therefore, our result can also infer an important insight that, though blood group AB has the highest risk of infection, it may not have high risk of severity. The large number of hospital data will be able to testify it further.

It has been shown recently that the Rh negative blood type has a protective role against SARS-CoV-2 ^12^. We have tested this association and found out 0.333(95% CI 0.152-0.587) seroprevalence for Rh negative individuals, which is not significantly different than 0.425(95% CI 0.382-0.469) seroprevalence of Rh positive people (p>0.05) (Supplementary Table 2). Thus, we did not find any association of Rh factor with COVID-19. However, the limited sample size (15), of Rh negative individuals should be taken with caution.

In summary, this is the first study in our knowledge which has been done on association of ABO locus with the COVID-19 susceptibility in India using asymptomatic COVID-19 positive individuals. Consistent with the previous observations, we have also found that the blood group O has least risk. However, we did not find any higher risk for blood group A as reported earlier, rather we see severalfold increased risk of infection for blood group AB. With our novel sampling methodology, we have also able to deduce that though blood group AB is highly susceptible for SARS-CoV-2 infection, it is unlike for them to develop disease severity.

## Data Availability

All the data is available with the manuscript.

## Acknowledgements

We are grateful to the Biosense Technologies, India for their kind help in the seroprevalence project. We did not receive any specific funding for this work.

## Author Contributions

GC conceived and designed this study. PPS, AKS, AS, PG, SM, SU, RKP, ANS, PD, VS, RM, MS, GOC, PK, VR, YBT, AP, VNM, CBM, PS and GC have collected the field data and performed the antibody as well as blood group testing. PPS and GC analysed the data. PPS, AKS, AS, PG, CBM and GC wrote the manuscript from the inputs of all co-authors. All authors contributed to the article and approved the submitted version.

## Data Availability Statement

All datasets generated for this study are included in the article/Supplementary Material.

## Figure Legends

**Supplementary Figure 1.**

The sampling locations in the Eastern Uttar Pradesh.

**Supplementary Figure 2.**

The sample test of seropositive and blood group of an individual.

**Supplementary Figure 3.**

The comparative bar-plot of various blood groups observed in present study as well as collected from published data. The error bars have been calculated at 95% confidence interval.

**Supplementary Table 1.** The distribution of ABO blood groups among various cohorts used in this study

|  | N | A+ | B+ | O+ | AB+ | A- | B- | O- |
| --- | --- | --- | --- | --- | --- | --- | --- | --- |
| All Samples | 509 | 0.198 (0.166-0.235) | 0.34 (0.3-0.382) | 0.269 (0.232-0.309) | 0.163 (0.134-0.198) | 0.006 (0.002-0.017) | 0.014 (0.007-0.028) | 0.010 (0.004-0.023) |
| Seropositive | 215 | 0.214 (0.164-0.274) | 0.298 (0.241-0.362) | 0.107 (0.073-0.155) | 0.358 (0.297-0.424) | 0.009 (0.003-0.033) | 0.014 (0.005-0.04) | 0 (0-0.017) |
|  | N | A | B | O | AB |  |  |  |
| All Samples | 509 | 0.204 (0.172-0.242) | 0.354 (0.313-0.396) | 0.279 (0.242-0.320) | 0.163 (0.134-0.198) |  |  |  |
| Seropositive | 215 | 0.223 (0.173-0.284) | 0.312 (0.253-0.376) | 0.107 (0.073-0.155) | 0.358 (0.297-0.424) |  |  |  |
| Published Samples | 2272 | 0.239 (0.222-0.257) | 0.375 (0.356-0.396) | 0.297 (0.278-0.316) | 0.089 (0.078-0.101) |  |  |  |

**Supplementary Table 2.**
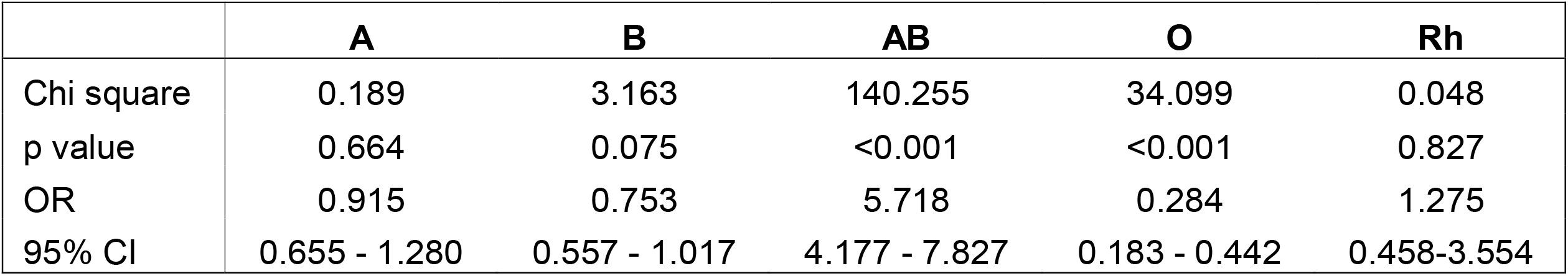
Various statistical tests on our and published data to infer the association and risk of ABO blood group for COVID-19. OR= Odds Ratio, CI= Confidence interval.

|  | **A** | **B** | **AB** | **O** | **Rh** |
| --- | --- | --- | --- | --- | --- |
| Chi square | 0.189 | 3.163 | 140.255 | 34.099 | 0.048 |
| p value | 0.664 | 0.075 | <0.001 | <0.001 | 0.827 |
| OR | 0.915 | 0.753 | 5.718 | 0.284 | 1.275 |
| 95% CI | 0.655 - 1.280 | 0.557 - 1.017 | 4.177 - 7.827 | 0.183 - 0.442 | 0.458-3.554 |

