## Supplementary figures and images for "The association of ABO blood group with the asymptomatic COVID-19 cases in India"

### Supplementary Figure 1

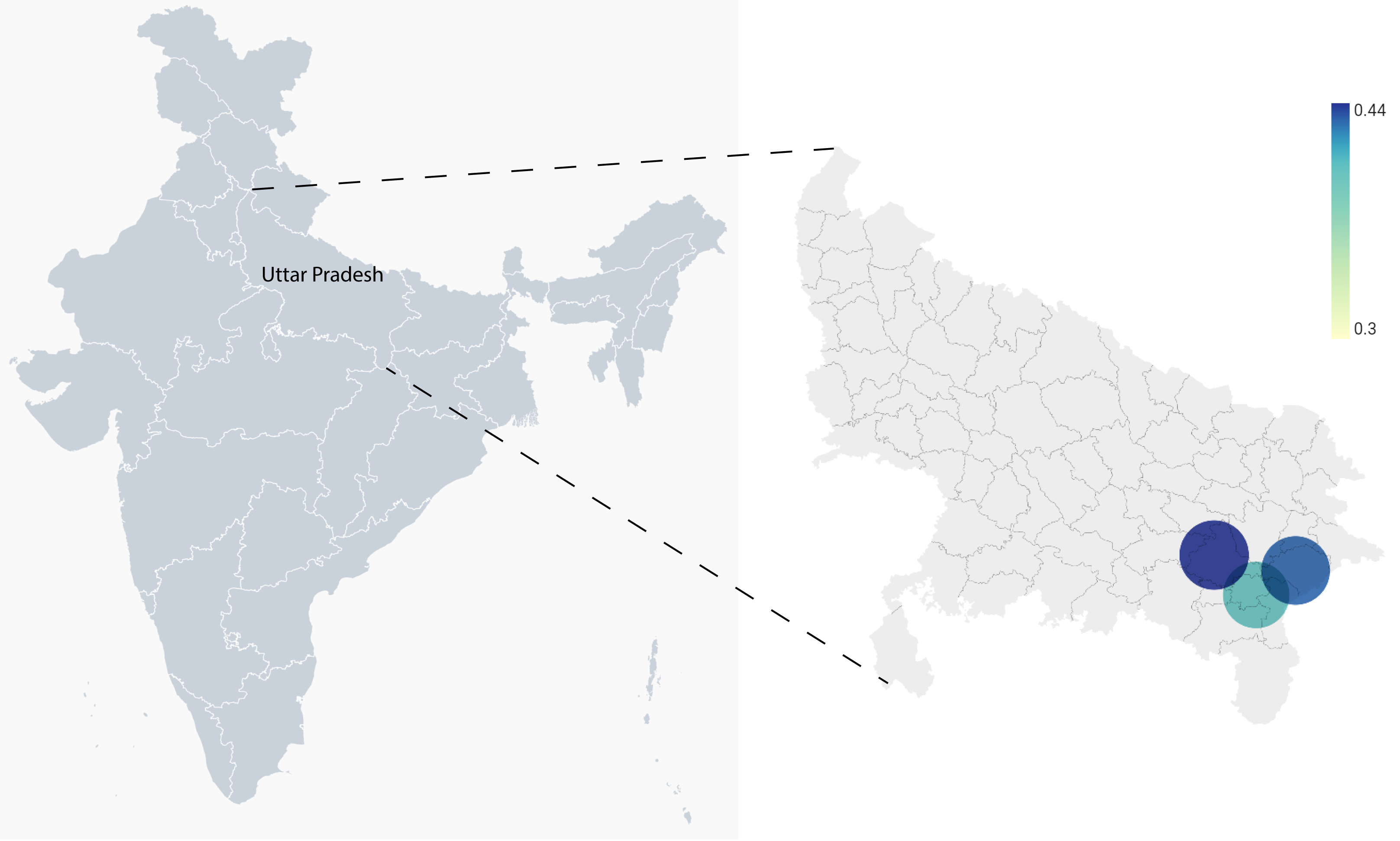

### Supplementary Figure 2

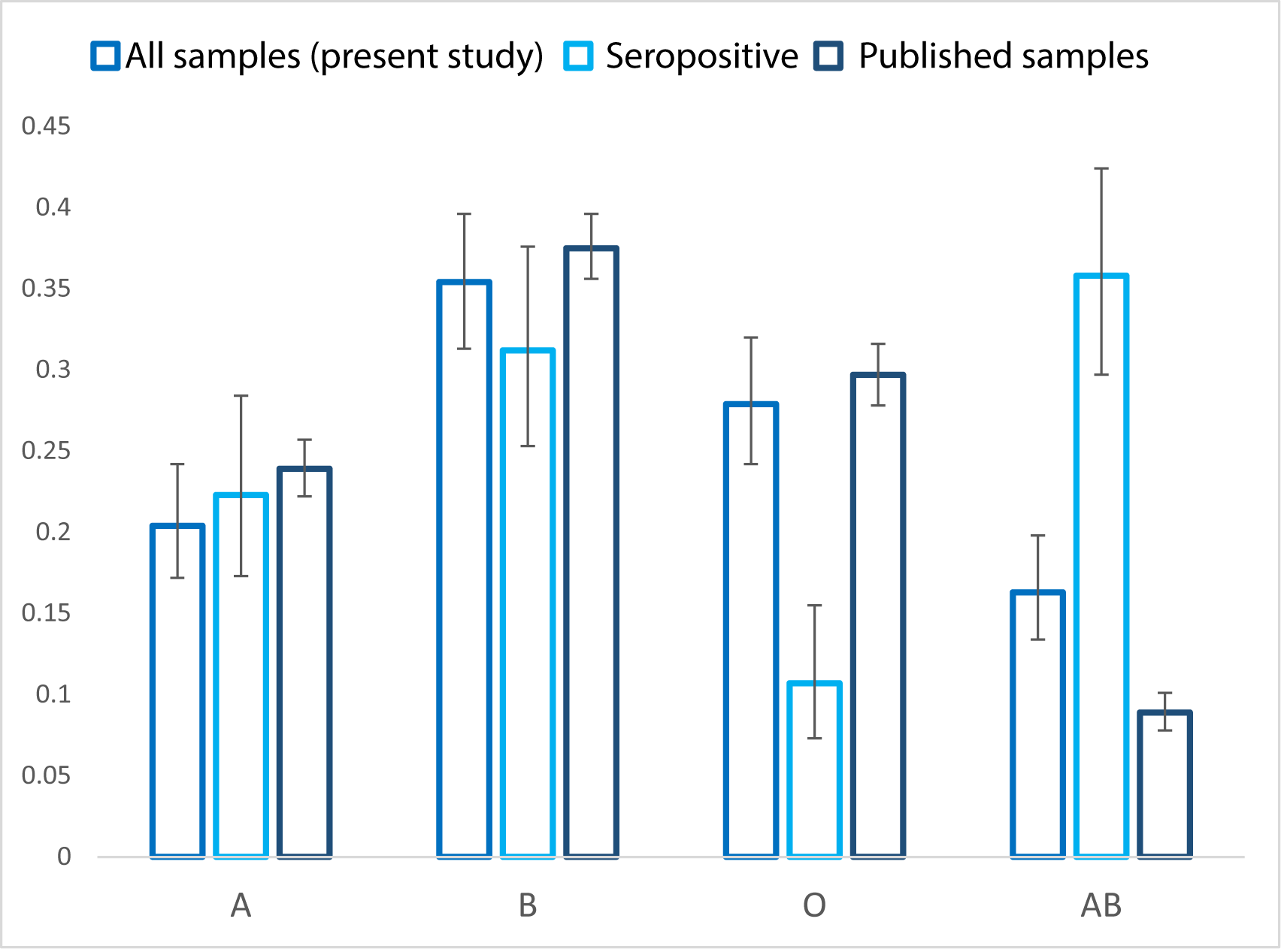
